# One year health outcomes associated with systemic corticosteroids for COVID-19: a longitudinal cohort study

**DOI:** 10.1101/2023.11.09.23298162

**Authors:** Olivia C Leavy, Richard J Russell, Ewen M Harrison, Nazir I Lone, Steven Kerr, Annemarie B Docherty, Aziz Sheikh, Matthew Richardson, Omer Elneima, Neil J Greening, Victoria Claire Harris, Linzy Houchen-Wolloff, Hamish J C McAuley, Ruth M Saunders, Marco Sereno, Aarti Shikotra, Amisha Singapuri, Raminder Aul, Paul Beirne, Charlotte E Bolton, Jeremy S Brown, Gourab Choudhury, Nawar Diar Bakerly, Nicholas Easom, Carlos Echevarria, Jonathan Fuld, Nick Hart, John R Hurst, Mark Jones, Dhruv Parekh, Paul Pfeffer, Najib M Rahman, Sarah Rowland-Jones, Ajay M Shah, Dan G Wootton, Caroline Jolley, AA Roger Thompson, Trudie Chalder, Melanie J Davies, Anthony De Soyza, John R Geddes, William Greenhalf, Simon Heller, Luke Howard, Joseph Jacob, R Gisli Jenkins, Janet M Lord, Will D-C Man, Gerry P McCann, Stefan Neubauer, Peter JM Openshaw, Joanna Porter, Matthew J Rowland, Janet T Scott, Malcolm G Semple, Sally J Singh, David Thomas, Mark Toshner, Keir Lewis, Liam G Heaney, Andrew Briggs, Bang Zheng, Mathew Thorpe, Jennifer K Quint, James D Chalmers, Ling-Pei Ho, Alex Horsley, Michael Marks, Krisnah Poinasamy, Betty Raman, Louise V Wain, Christopher E Brightling, Rachael A Evans, the PHOSP-COVID Collaborative Group

## Abstract

**Background:** In patients with COVID-19 requiring supplemental oxygen, dexamethasone reduces acute severity and improves survival, but longer-term effects are unknown. We hypothesised that systemic corticosteroid administration during acute COVID-19 would be associated with improved health-related quality of life (HRQoL) one year after discharge.

**Methods:** Adults admitted to hospital between February 2020 and March 2021 for COVID-19 and meeting current guideline recommendations for dexamethasone treatment were included using two prospective UK cohort studies. HRQoL, assessed by EQ-5D-5L utility index, pre-hospital and one year after discharge were compared between those receiving corticosteroids or not after propensity weighting for treatment. Secondary outcomes included patient reported recovery, physical and mental health status, and measures of organ impairment. Sensitivity analyses were undertaken to account for survival and selection bias.

**Findings:** In 1,888 participants included in the primary analysis, 1,149 received corticosteroids. There was no between-group difference in EQ-5D-5L utility index at one year (mean difference 0.004, 95% CI: -0.026 to 0.034, *p* = 0.77). A similar reduction in EQ-5D-5L was seen at one year between corticosteroid exposed and non-exposed groups (mean (SD) change -0.12 (0.22) vs -0.11 (0.22), *p* = 0.32). Overall, there were no differences in secondary outcome measures. After sensitivity analyses modelled using a larger cohort of 109,318 patients admitted to hospital with COVID-19, EQ-5D-5L utility index at one year remained similar between the two groups.

**Interpretation:** Systemic corticosteroids for acute COVID-19 have no impact on the large reduction in HRQoL one year after hospital discharge. Treatments to address this are urgently needed.

**Take home message:** Systemic corticosteroids given for acute COVID-19 do not affect health-related quality of life or other patient reported outcomes, physical and mental health outcomes, and organ function one year after hospital discharge

## Introduction

The discovery of vaccines and effective treatments for acute COVID-19 (corticosteroids [dexamethasone], anti-interleukin (IL)-6 agents, monoclonal antibodies and Janus kinase inhibitors) have reduced progression to invasive mechanical ventilation and improved mortality [1-4]. However, many survivors experience persistent symptoms, physical and mental health effects, cognitive impairment, and multi-system organ damage, which can reduce health-related quality of life (HRQoL) for years after the initial infection [5-7].

Definitions for post-COVID-19 sequelae vary [8, 9], but the patient-derived term ‘Long Covid’ is now commonly used to describe persistent symptoms beyond four weeks after the acute infection [10]. The mechanisms underlying Long Covid are complex, multifaceted, and not yet fully understood, but potentially include persistent inflammation, which is associated with the severity of ongoing health impairments [5, 11]. Corticosteroids prescribed for acute COVID-19 may potentially reduce the risk and severity of Long Covid by attenuating the acute inflammatory burden.

Many of the large acute COVID-19 therapeutic trials including RECOVERY [1-4] did not have detailed follow-up, which limits understanding of the longer-term effects, and it would now be unethical to randomise patients to placebo rather than corticosteroids. Adults previously randomised to receive acute corticosteroids on intensive care showed no improvement in HRQoL at six months compared to usual care [12], and we previously reported no acute corticosteroid effect on patient perceived recovery at one year[6]. However, it is unknown whether corticosteroids during acute COVID-19 affect other longer-term sequelae.

Using data from the PHOSP-COVID (Post-hospitalisation COVID-19 study) [13] and ISARIC (International Severe Acute Respiratory and emerging Infection Collaboration) [14] studies, we aimed to investigate whether treatment with corticosteroids in patients with COVID-19 requiring oxygen supplementation was associated with improved HRQoL one year after hospital discharge. Additionally, we aimed to investigate the effect of acute corticosteroids on a broad range of secondary health outcomes.

## Methods

### Study design

This was a longitudinal cohort study using data from two UK multicentre prospective cohort studies. Adults discharged from hospital after COVID-19 between 1^st^ February 2020 and 31^st^ March 2021 were recruited from 36 UK National Health Service (NHS) hospital sites as part of the PHOSP-COVID study previously described [13]. Data were collected one year after hospital discharge, including patient reported recovery, physical and mental health status, and measures of organ impairment (detailed below). Pre-hospital EuroQol five-dimension five-level utility index (EQ-5D-5L UI) was completed retrospectively at a study visit 2-7 months after hospital discharge, with participants considering their quality of life prior to admission for COVID-19.

For the sensitivity analysis, we used data from the ISARIC (UK) study [14], which included more than 300,000 patients admitted to over 200 NHS hospitals across England, Scotland and Wales with COVID-19.

### Participants

Eligibility criteria for PHOSP-COVID have been previously described in detail [13]. For this analysis we selected participants who required supplemental oxygen therapy (WHO clinical progression scale 5), non-invasive ventilatory support (WHO clinical progression scale 6), or invasive mechanical ventilation (WHO clinical progression scale 7-9) [15] during their hospital admission in accordance with current guideline requirements for corticosteroid use in COVID-19 [16], and who had completed an EQ-5D-5L UI at their one year study visit. We excluded patients on pre-existing immunosuppressant medications, and where corticosteroid exposure was unknown or not recorded (Figure 1).

**Figure 1:**
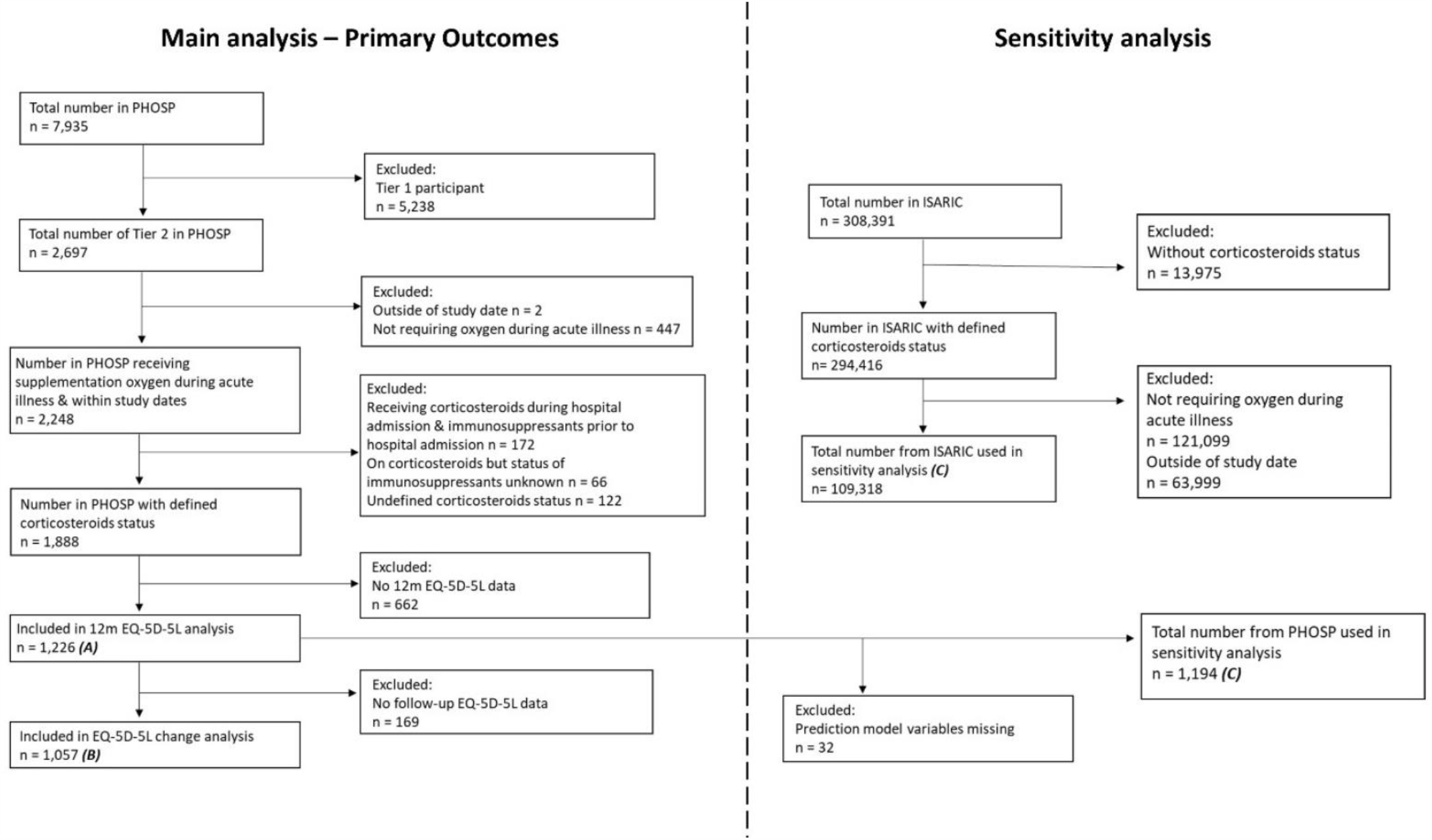
Consort diagram demonstrating study population included in co-primary outcomes of i) EQ-5D-5L UI at one year (A) and ii) change in EQ-5D-5L UI from pre-hospital to one year (B), and sensitivity analysis (C).

For the sensitivity analysis, we analysed a subset of the ISARIC study cohort, who were admitted with COVID-19 in the same study period and meeting the same WHO clinical progression scale criteria [14] (Figure 1).

### Exposure

Patients who received any systemic (oral or intravenous) corticosteroid during their hospital admission for COVID-19 were compared to those who did not.

### Outcomes

The primary outcome was HRQoL, assessed by EQ-5D-5L UI [17]. EQ-5D-5L one year after hospital discharge, and change in EQ-5D-5L UI from pre-hospital to one year, were compared between corticosteroid exposed and non-exposed patients.

Secondary outcomes were patient perceived recovery (patient reported recovery rate, symptom count, fatigue visual analogue scale (VAS), breathlessness VAS), physical health status (dyspnoea-12 score [18], Functional Assessment of Chronic Illness Therapy (FACIT) fatigue score [19], Washington Group Short Set on Functioning (WG-SS) score [20], incremental shuttle walk test (ISWT) distance [21], short physical performance battery (SPPB) score [22]), mental health status (Montreal Cognitive Assessment (MoCA) score [23], Generalised Anxiety Disorder (GAD)-7 score [24], Patient Health Questionnaire (PHQ)-8 score [25], Post-traumatic Stress Disorder Checklist (PCL)-5 score [26]), and organ function (forced expiratory volume in 1 second (FEV1), forced vital capacity (FVC), FEV1/FVC ratio, carbon monoxide transfer coefficient (KCO), transfer factor of the lung for carbon monoxide (TLCO), brain-natriuretic peptide (BNP), haemoglobin A1c (HbA1c), estimated glomerular filtration rate (eGFR), C-reactive protein (CRP), fibrinogen).

### Bias

Several potential sources of bias were considered *a priori*; (i) *treatment bias* by clinician prescribing decision (prior to corticosteroids becoming standard care in June 2020), (ii) *selection bias* regarding who participated in the PHOSP-COVID study, and (iii) *survivor bias* due to participants being recruited to PHOSP-COVID after hospital discharge (i.e., survivors). A statistical analysis plan was developed including the use of propensity weighting to ensure balance between treatment groups in the primary analysis, and sensitivity analyses using data from the ISARIC study.

### Statistical analysis

The main analysis was undertaken using the PHOSP-COVID cohort. A logistic regression model was fitted to estimate propensity for exposure to corticosteroids. An average treatment effect of corticosteroid treatment on the outcomes (primary and secondary) was calculated weighted by the inverse of propensity for exposure using either linear or logistic regression, depending on the distribution of the outcome. The following variables, which potentially influence treatment decisions, were included in the propensity model: age, sex, obesity status, ethnicity, Index of Multiple Deprivation, WHO Clinical Progression Scale status, smoking status, presence of specific comorbidities (cardiovascular, respiratory, metabolic/endocrine/renal, neurological/psychiatric) and total number of comorbidities. Multiple Imputation by Chained Equations (MICE) was performed to deal with missing data for the variables used in the propensity model. Summary statistics tables were produced for patients by exposure status, visually inspecting the distribution of propensity scores and evaluating imbalance between groups by standardised mean difference (SMD).

### Sensitivity analyses

Sensitivity analyses were performed using the ISARIC (UK) dataset to address selection, treatment and survivor biases in PHOSP-COVID (supplementary methods). In summary, a propensity score weighting for corticosteroid treatment was developed in the ISARIC (UK) cohort (survivors and non-survivors) using logistic regression. The PHOSP-COVID dataset was used to develop a prediction model for EQ-5D-5L UI at one year. We used this model to calculate predicted one-year EQ-5D-5L UI values for those that survived COVID-19 hospitalisation in the ISARIC cohort (1000 estimates per patient). Adults that did not survive were assigned an EQ-5D-5L value of zero. Participants who were in both ISARIC and PHOSP-COVID cohorts were assigned their PHOSP-COVID EQ-5D-5L value. The 1000 datasets created were sub-sampled down to the PHOSP-COVID dataset size to ensure robust standard errors (1000 random samples of each dataset). These datasets were used to produce an average treatment effect of corticosteroid exposure on EQ-5D-5L UI weighted by the inverse of propensity for exposure using linear regression.

The sensitivity analysis addressed selection and survivor bias by using the structure of the ISARIC population (assuming the ISARIC population was similar to all hospitalised patients with COVID-19 eligibility to receive corticosteroids). The ISARIC cohort included participants who did not survive hospitalisation with COVID-19. Biased treatment assignment was accounted for by developing a propensity score with corticosteroid as the dependent variable, which was developed in the ISARIC cohort, and was therefore independent of survival status at hospital discharge.

Statistical analysis was undertaken using R (version 4.2.0) with the *tidyverse, tidymodels, mice, finalfit, WeightIt, and tableone* packages for all statistical analyses. The study is reported using the STROBE reporting guidelines.

### Permissions

PHOSP-COVID was approved by the Leeds West Research Ethics Committee (20/YH/0225) and is registered on the ISRCTN Registry (ISRCTN10980107). ISARIC was approved by the South Central -Oxford C Research Ethics Committee in England and the Scotland A Research Ethics Committee.

## Results

The relevant PHOSP-COVID cohort consisted of 2,697 participants, of whom 2,248 required at least supplemental oxygen and were discharged from hospital between 1^st^ February 2020 and 31^st^ March 2021. There were 1,888 participants with non-missing corticosteroid information not prescribed immunosuppressant medication pre-hospital, of which 1,149 (60.9%) were corticosteroid exposed, and 739 (39.1%) were corticosteroid non-exposed. 1,226 participants had an EQ-5D-5L UI score at their one-year visit and 1,057 participants had both pre-hospital and one-year EQ-5D-5L UI scores (Figure 1).

Baseline characteristics for the 1,888 included participants demonstrated a mean age of 58.6 years with 64.4% being male. 75.1% were White, 10.1% South Asian, 7.3% Black, and 7.5% other ethnicity. 58.6% were obese (body mass index ≥30kg/m^2^), and 43.8% had two or more comorbidities (Table S1). Prior to propensity weighting some baseline characteristics were imbalanced between treatment groups, as demonstrated by an SMD of >0.1 (Table S1). Participants treated with corticosteroids were slightly younger compared to those not receiving corticosteroids (58.0 vs 59.7 years), and had greater prevalence of: White ethnicity (76.8% vs 72.5%), deprivation (49.5% vs 41.0% in lowest two deprivation index quintiles), and obesity (61.0% vs 55.8%). The corticosteroid group had a lower proportion of ‘never smokers’ (54.9% vs 56.4%). There were also differences in the level of respiratory support required between patients treated with corticosteroid and those not: 51.5% vs 54.7% received low-flow oxygen (WHO scale 5), 33.3% vs 22.6% received non-invasive respiratory support (WHO scale 6), 15.1% vs 22.7% received invasive mechanical ventilation (WHO scale 7-9).

Propensity weighting successfully achieved balance between the treatment groups, as demonstrated by a SMD <0.1 for all recorded baseline outcomes (Table 1).

**Table 1:**
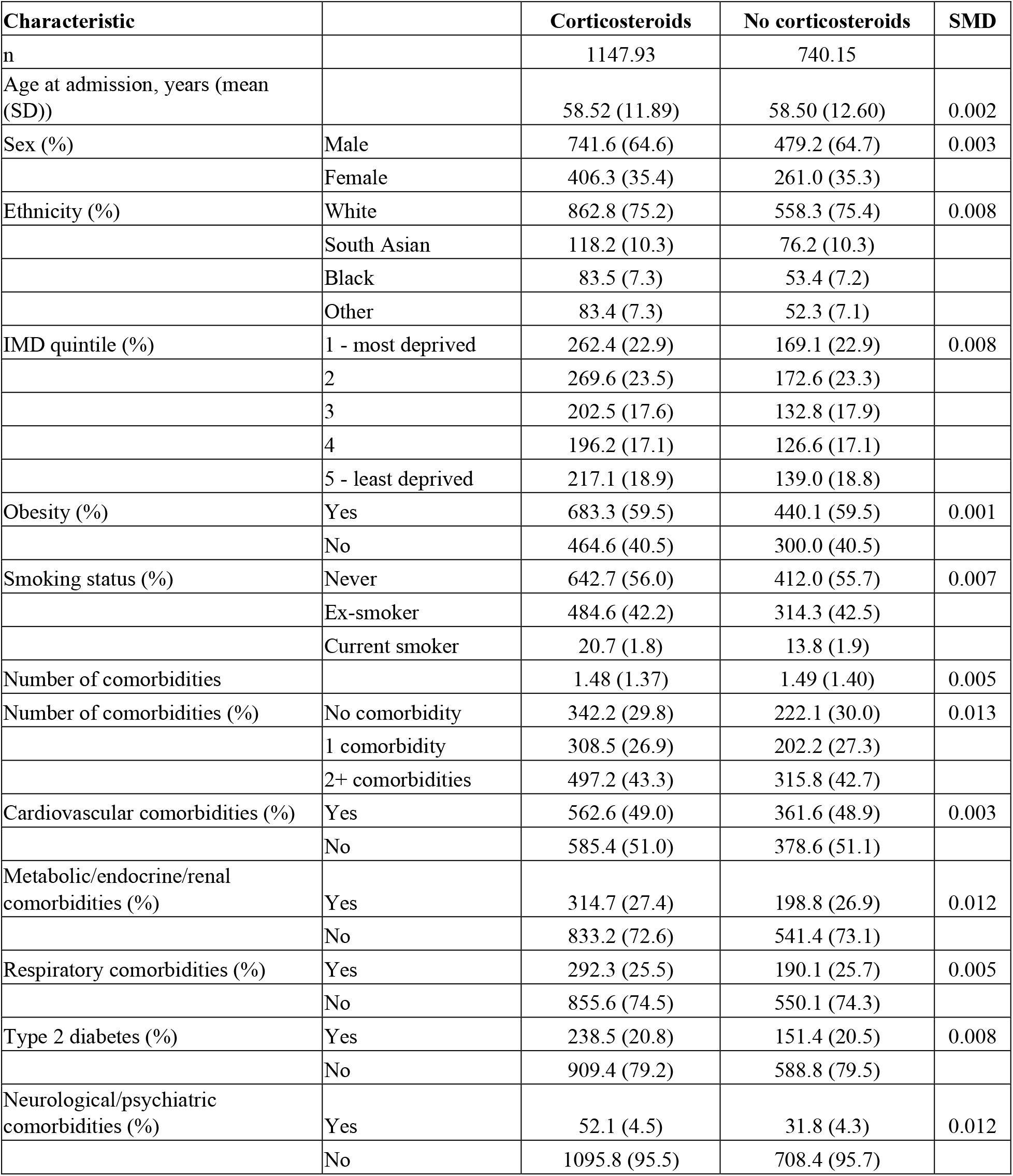
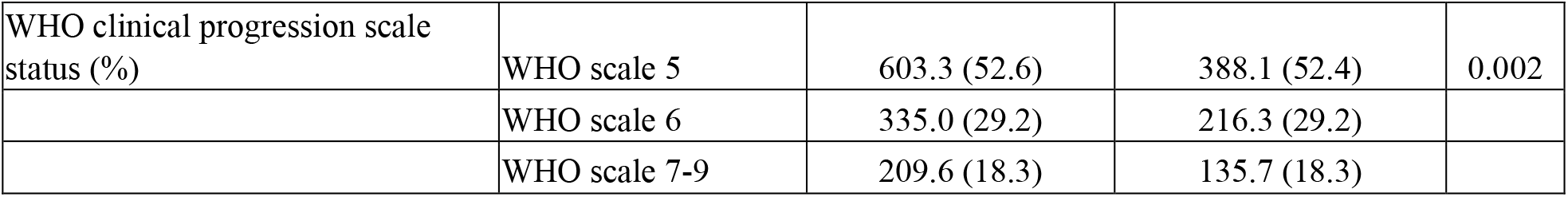
Baseline characteristics after propensity weighting. Data are n, n (%) or mean (SD). Percentages are calculated by category after exclusion of missing data for that variable. SD = standard deviation. IMD = Index of Multiple Deprivation. WHO = World Health Organisation. SMD = Standardised Mean Difference

### Primary outcomes

After propensity weighting for treatment there was no statistically significant difference in EQ-5D-5L UI at one year between corticosteroid exposed (mean (SD) 0.72 (0.25)) and non-exposed (0.71 (0.25)) groups (mean difference 0.004, 95% CI: -0.026 to 0.034, *p* = 0.77) (Table 2 and Figures 2 and 3).

**Table 2:** Primary and secondary outcomes: patient reported outcomes, mental health status and cognitive assessments. Data are n, n (%), mean (SD) or median (IQR). Percentages are calculated by category after exclusion of missing data for that variable. SD = standard deviation, IQR = interquartile range, EQ-5D-5L UI = EuroQol-5-Dimension 5-level utility index, VAS = visual analogue scale, FACIT = Functional Assessment of Chronic Illness Therapy, MoCA = Montreal Cognitive Assessment, WG-SS = Washington Group Short Set, GAD-7 = Generalized Anxiety Disorder 7-item score, PHQ-8 = Patient Health Questionnaire-8, PCL-5 = Post-Traumatic Stress Disorder Checklist for the Diagnostic and Statistical Manual of Mental Disorders, PTSD = post-traumatic stress disorder

| Outcome |  | Corticosteroids | No corticosteroids | p-value |
| --- | --- | --- | --- | --- |
| n |  | 737.5 | 488.9 |  |
| EQ-5D-5L UI at 1 year (mean (SD)) |  | 0.72 (0.25) | 0.71 (0.25) | 0.773 |
| EQ-5D-5L UI change pre-hospital to 1 year (mean (SD)) |  | -0.11 (0.22) | -0.12 (0.22) | 0.317 |
| Do you feel fully recovered from covid? (n (%)) | Yes | 223.1 (30.2) | 139.1 (28.5) | 0.811 |
|  | No/not sure | 465.3 (63.1) | 299.6 (61.3) |  |
|  | Missing | 49.2 (6.7) | 50.2 (10.3) |  |
| Any symptom at 1 year (n (%)) | Yes | 656.8 (89.1) | 423.6 (86.6) | 0.508 |
|  | No | 39.9 (5.4) | 21.4 (4.4) |  |
|  | Missing | 40.8 (5.5) | 43.9 (9.0) |  |
| Symptom count (median [IQR]) |  | 8.00 [4.00, 16.00] | 9.00 [4.00, 16.00] | 0.671 |
| Fatigue VAS |  | 2.00 [0.00, 5.00] | 3.00 [0.00, 5.00] | 0.465 |
| Breathlessness VAS |  | 1.00 [0.00, 4.00] | 1.00 [0.00, 5.00] | 0.043 |
| Dyspnoea-12 score (mean (SD)) |  | 5.04 (7.22) | 5.46 (7.83) | 0.373 |
| FACIT fatigue score (mean (SD)) |  | 36.79 (12.23) | 36.31 (12.87) | 0.524 |
| MoCA corrected (mean (SD)) |  | 26.90 (3.22) | 26.65 (3.23) | 0.232 |
| MoCA corrected <23 (n (%)) | Yes | 49.6 (6.7) | 41.7 (8.5) | 0.373 |
|  | No | 516.8 (70.1) | 356.6 (72.9) |  |
|  | Missing | 171.1 (23.2) | 90.6 (18.5) |  |
| WG-SS score (median [IQR]) |  | 2.00 [0.00, 4.00] | 2.00 [0.00, 4.00] | 0.613 |
| GAD-7 total score (mean (SD)) |  | 4.75 (5.46) | 4.91 (5.60) | 0.631 |
| Anxiety (GAD-7 score >8) (n (%)) | Yes | 159.6 (21.6) | 110.5 (22.6) | 0.684 |
|  | No | 576.0 (78.1) | 375.8 (76.9) |  |
|  | Missing | < 5 | < 5 |  |
| PHQ-8 total score (mean (SD)) |  | 6.14 (6.25) | 6.21 (6.39) | 0.791 |
| PCL-5 total score (mean (SD)) |  | 13.63 (16.76) | 13.76 (17.54) | 0.901 |
| PTSD (PCL-5 score ≥38) (n (%)) | Yes | 79.6 (10.8) | 51.0 (10.4) | 0.866 |
|  | No | 650.8 (88.2) | 431.1 (88.2) |  |
|  | Missing | 7.1 (1.0) | 6.7 (1.4) |  |

**Figure 2:**
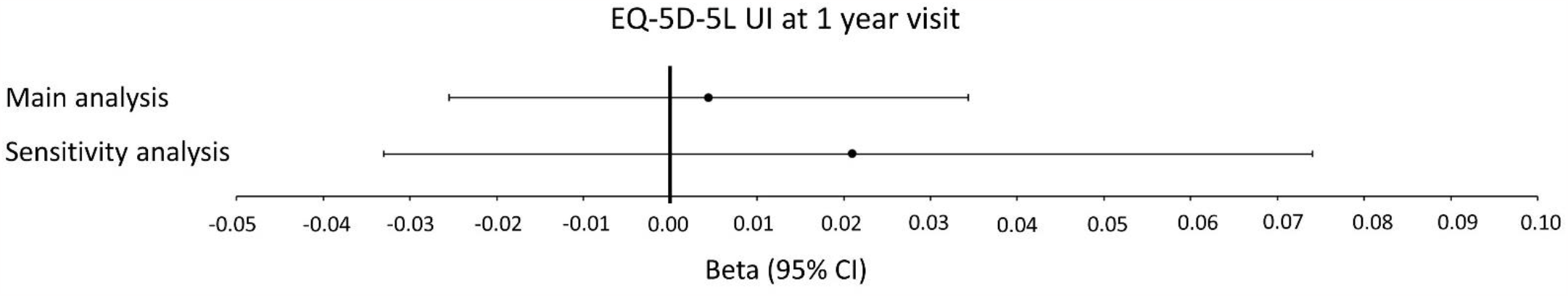
EQ-5D-5L UI at one year after hospital discharge in corticosteroid exposed vs non-exposed patients.

**Figure 3:**
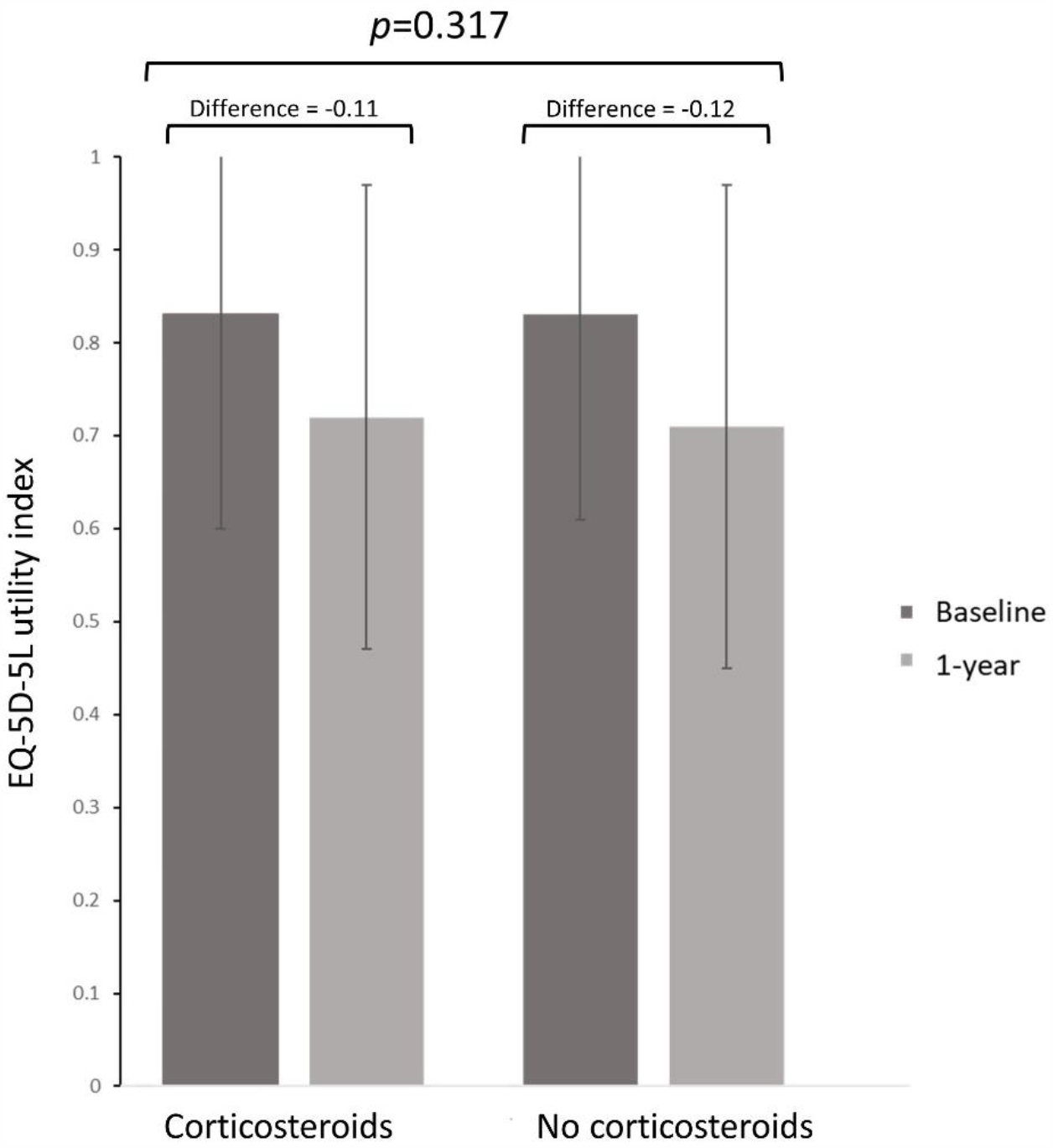
EQ-5D-5L UI change from pre-hospital (baseline) to one year in corticosteroid exposed vs non-exposed patients.

There was a large reduction in EQ-5D-5L UI from pre-hospital to one year, with no significant difference between corticosteroid exposed (mean change -0.12 (0.22)) and non-exposed (−0.11 (0.22)) groups (mean difference 0.01, 95% CI: -0.01 to 0.04, *p* = 0.32) (Table 2 and Figure 3).

### Secondary outcomes

Secondary outcomes, assessing patient reported outcomes, physical, cognitive and mental health status, and measurements of organ impairment, were not significantly different between treatment groups at one year (Tables 2 and 3, and Figure 4), except breathlessness VAS which was lower in patients who had received corticosteroids (median [IQR] 1.0 [0.0, 4.0] vs 1.0 [0.0, 5.0], *p* = 0.043).

**Table 3:** Secondary outcomes: physical impairment and organ function. Data are n, n (%) or mean (SD). Percentages are calculated by category after exclusion of missing data for that variable. SD = standard deviation, ISWT = incremental shuttle walk test, SPPB = short physical performance battery, FEV1 = forced expiratory volume, FVC = forced vital capacity, KCO = carbon monoxide transfer coefficient, TLCO = transfer capacity of the lung, BNP = brain natriuretic peptide, eGFR = estimated glomerular filtration rate

| Outcome |  | Corticosteroids | No corticosteroids | P-value |
| --- | --- | --- | --- | --- |
| n |  | 737.5 | 488.9 |  |
| ISWT distance, m (mean (SD)) |  | 462.78 (468.30) | 455.15 (252.13) | 0.770 |
| ISWT % predicted (mean (SD)) |  | 62.59 (59.25) | 60.59 (28.72) | 0.604 |
| SPPB total score (mean (SD)) |  | 10.12 (1.99) | 9.93 (2.32) | 0.160 |
| SPPB (mobility disability $\leq 10$) (n (%)) | Yes | 289.4 (39.2) | 219.0 (44.8) | 0.671 |
|  | No | 325.1 (44.1) | 233.1 (47.7) |  |
|  | Missing | 122.9 (16.7) | 36.8 (7.5) |  |
| FEV1 % predicted $< 80\%$ (n (%)) | Yes | 78.1 (10.6) | 70.3 (14.4) | 0.613 |
|  | No | 258.1 (35.0) | 210.6 (43.1) |  |
|  | Missing | 401.3 (54.4) | 208.0 (42.5) |  |
| FVC % predicted $< 80\%$ (n (%)) | Yes | 85.1 (11.5) | 74.3 (15.2) | 0.772 |
|  | No | 251.1 (34.0) | 207.3 (42.4) |  |
|  | Missing | 401.3 (54.4) | 207.3 (42.4) |  |
| FEV1/FVC $< 0.7$ (n (%)) | Yes | 32.5 (4.4) | 31.1 (6.4) | 0.606 |
|  | No | 310.6 (42.1) | 257.8 (52.7) |  |
|  | Missing | 394.4 (53.5) | 200.0 (40.9) |  |
| KCO $< 80\%$ predicted (n (%)) | Yes | 15.6 (2.1) | 9.3 (1.9) | 0.287 |
|  | No | 103.6 (14.0) | 98.0 (20.1) |  |
|  | Missing | 618.3 (83.8) | 381.6 (78.0) |  |
| TLCO $< 80\%$ predicted (n (%)) | Yes | 19.2 (2.6) | 27.7 (5.7) | 0.074 |
|  | No | 92.4 (12.5) | 71.4 (14.6) |  |
|  | Missing | 625.8 (84.9) | 389.8 (79.7) |  |
| BNP $\geq 100$ ng/L or pro-NT-BNP $\geq 400$ ng/L | Yes | 23 (3.2) | 24 (4.9) | 0.529 |
|  | No | 292 (39.9) | 229 (46.3) |  |
|  | Missing | 416 (56.9) | 242 (48.9) |  |
| HbA1C $\geq 6.0\%$ (DCCT/NGSP) | Yes | 157 (21.5) | 114 (23.0) | 0.881 |
|  | No | 277 (37.9) | 196 (39.6) |  |
|  | Missing | 297 (40.6) | 185 (37.4) |  |
| eGFR $< 60$ mL/min per $1.73 \text{ m}^2$ | Yes | 74 (10.1) | 63 (12.7) | 0.586 |
|  | No | 475 (65.0) | 321 (64.8) |  |
|  | Missing | 182 (24.9) | 111 (22.4) |  |
| C-reactive protein concentration $> 5$ mg/L | Yes | 124 (17.0) | 78 (15.8) | 0.204 |
|  | No | 423 (57.9) | 321 (64.8) |  |
|  | Missing | 184 (25.2) | 96 (19.4) |  |
| Fibrinogen (g/L) (mean (SD)) |  | 3.58 (2.23) | 3.56 (0.87) | 0.846 |

**Figure 4:**
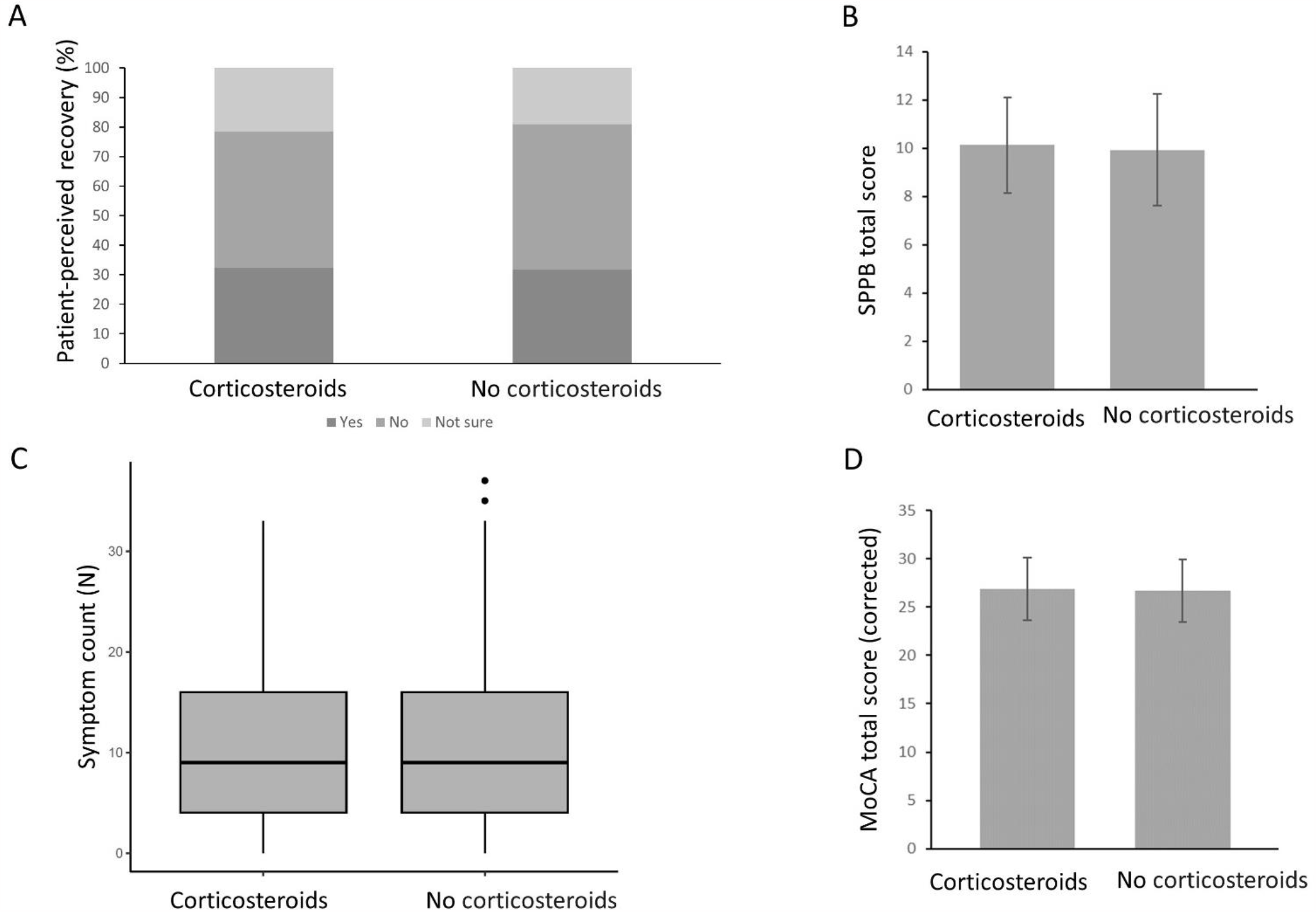
Secondary outcomes: patient-perceived recovery (A), Short Physical Performance Battery score (B), symptom count (C) and Montreal Cognitive Assessment score (corrected) (D) one year after hospital discharge in corticosteroid exposed vs non-exposed patients.

### Sensitivity analysis

In the sensitivity analysis, there was no significant difference in the EQ-5D-5L at one year between patients who received corticosteroids and those who did not (between group difference 0.021, 95% CI: -0.033 to 0.074, *p* = 0.45) (Figure 2).

## Discussion

To our knowledge this is the first report investigating the effect of acute corticosteroids on HRQoL, other patient reported outcomes, physical and mental health status, and multi-system organ effects one year after hospitalisation for COVID-19. We observed large reductions in HRQoL at one year and report novel findings that there was neither a difference in EQ-5D-5L UI at one year, nor in EQ-5D-5L change pre-hospital to one year, between patients who did or did not receive corticosteroids for their acute illness. There remained no difference in HRQoL at one year after adjusting for survivor and selection bias using a large cohort of patients admitted with COVID-19 (ISARIC cohort). We also found no difference between receipt of acute corticosteroids or not across a range of secondary endpoints assessing patient reported outcomes, physical, cognitive and mental health status, and measurements of multi-system organ impairment. Despite the observational longitudinal nature of our study, it is likely to be the most comprehensive and robust data available, as the large acute randomised controlled trials of therapeutics in COVID-19 were unable to perform in-person follow-up assessments [1-4], and corticosteroids are now standard of care for COVID-19, meaning a placebo-controlled trial would now be unethical [16]. Our recruitment period encompassed time before and after systemic corticosteroids became standard care for patients requiring oxygen due to COVID-19 (June 2020), allowing comparison between corticosteroid exposed and non-exposed groups.

Our data demonstrate the significant negative impact on HRQoL and other health outcomes one year after hospital discharge in this population, similar to our previous reports but in a larger subset [6]. Pre-hospital our cohort reported EQ-5D-5L UI scores in line with normal values (reported as 0.81 for men and 0.79 for women aged 55-59 years) [27]. One year after discharge from hospital, the EQ-5D-5L UI was comparable to long-term health conditions such as COPD [28].

Developments in treatments for acute COVID-19 (including pharmacological therapies, such as corticosteroids, and ventilation strategies), combined with effective vaccines, have significantly reduced the risk of in-hospital COVID-19 mortality. However, the risk of Long Covid remains, and although risk increases with more severe acute illness [5], many people with mild acute COVID-19 develop persistent health problems. We have previously shown that elevated inflammatory proteins five months after COVID-19 hospital discharge are associated with increased risk of very severe health impairments at one year [5], therefore it was reasonable to hypothesize that the anti-inflammatory effect of corticosteroids could mitigate the risk of Long Covid. A previous study found no difference in HRQoL 180 days after hospital discharge from a higher 12mg dose of dexamethasone compared to the standard 6mg dose [29], but HRQoL comparisons between corticosteroid treated or not were not available.

Other acute pharmacological interventions have shown promising effects on the risk of Long Covid. Anti-IL6 (tocilizumab) improves HRQoL at six months in COVID-19 survivors admitted to intensive care [12], although whether this benefit applies to patients outside of intensive care is unknown. The anti-viral remdesivir is associated with a reduction in rates of Long Covid at 180 days, although the study excluded severely unwell patients so this benefit may not apply to a broader population [30]. *Post-hoc* analysis of nebulised interferon-beta-1a for COVID-19 showed reductions in fatigue/malaise and loss of taste or smell at 60-90 days compared to placebo, and further investigation is ongoing [31]. Metformin reduces the risk of Long Covid in non-hospitalised overweight and obese patients, although the effect in more severe disease is unknown [32]. While the results of these trials are encouraging, it is noteworthy that each has limitations to their applicability in a wider patient population, and none have provided strong enough evidence to change treatment guidelines with the aim of reducing Long Covid. The HEAL-COVID study reported no benefit from 2 weeks of anticoagulation (apixaban) on post-discharge mortality or hospital readmission but has not yet reported quality of life outcomes [33]. A second study arm investigating 12 months of atorvastatin is underway [34].

Trials of potential treatments for patients with persistent health problems beyond the acute COVID-19 illness are being undertaken, although are few in number. In a phase 2 placebo-controlled trial, 4 weeks of AXA1125 (an endogenous metabolic modulator comprised of five amino acids and N-acetylcysteine) improved fatigue scores in patients with persistent fatigue at least 12 weeks after COVID-19 [35]. The Stimulate-ICP (Symptoms, Trajectory, Inequalities and Management: Understanding Long-COVID to Address and Transform Existing Integrated Care Pathways) study will investigate the effect of antihistamine (famotidine/loratidine), anticoagulation (rivaroxaban), and anti-inflammatory (colchicine) medications on Long Covid recovery, in addition to interventions such as rehabilitation strategies [36]. The PHOSP-I study will investigate tocilizumab in patients with persistent symptoms at least 3 months after COVID-19 and evidence of persistent systemic inflammation [37]. Given the evidence for acute interventions not reducing Long Covid across a broad patient population, these trials and others are urgently needed to reduce post-covid sequelae including Long Covid.

Our study has a number of strengths. We included a large cohort of patients discharged from hospital after receiving oxygen for COVID-19, and our sensitivity analysis uses ISARIC data to verify our findings in a much larger hospitalised cohort also requiring oxygen. Therefore, we are confident that our findings are applicable to patients meeting guideline criteria for corticosteroid treatment for COVID-19. Additionally, we used propensity weighting to ensure balance between groups prior to analysing one-year outcomes, in an attempt to replicate the effect of randomised allocation and account for elements of biasing. We are confident, therefore, that the lack of benefit from acute corticosteroids observed here is genuine.

Our study has some limitations. First, despite using propensity weighting methods, this is an observational study and therefore unable to fully replicate a randomised trial. Our statistical methods were designed to minimise potential biases related to this, but some residual effect may remain. Second, we included patients admitted to hospital over a 14-month period, spanning waves of different covid variants, and the early stages of the vaccine rollout. We cannot exclude potential effects due to these factors, particularly as our corticosteroid non-exposed participants were predominantly hospitalised before June 2020, and corticosteroid exposed participants predominantly after June 2020. Third, the PHOSP-COVID cohort had a more severe acute illness than the general hospitalised COVID-19 population, and only includes patients who survived at least 5 months after discharge, and is therefore subject to selection and survivor biases. We have attempted to address these in our sensitivity analysis, using the ISARIC cohort which includes patients who died. Fourth, there is a significant amount of missing lung function data, due to variable infection prevention restrictions during the study period. Therefore, we cannot fully exclude a possible effect on lung function. Finally, pre-hospital EQ-5D-5L was assessed retrospectively using patient recollection of their quality of life prior to hospitalisation with COVID-19. These data are therefore subject to recall bias, although the effect is likely equal between the treatment groups.

There remains a large reduction in HRQoL, and other health outcomes, one year after hospitalisation for COVID-19. Studies to identify pharmacological and non-pharmacological interventions given after the acute COVID-19 illness are essential to address this. It is also important to seek better mechanistic understanding of post-covid sequelae and improve phenotyping of patients who may respond to specific interventions.

In conclusion, we found no long-term benefit on HRQoL from corticosteroids given to treat acute COVID-19. There remains an urgent need for effective interventions that reduce the long-term burden of health issues following COVID-19.

### Funding

PHOSP-COVID is supported by a grant from the MRC-UK Research and Innovation and the Department of Health and Social Care through the National Institute for Health Research (NIHR) rapid response panel to tackle COVID-19. The funder had no role in study design, data collection, data analysis, data interpretation, or writing of the report.

### Data access

The PHOSP-COVID study website (https://www.phosp.org) contains an overview of the study, resources, information about people involved, and publications. Research activity using the study is organised across a series of Working Groups. These were established at the outset of the study to coordinate research, minimise duplication of efforts, and facilitate communication across research and clinical specialties. Researchers interested in undertaking research using PHOSP-COVID are encouraged to contact the relevant Working Group leads (https://www.phosp.org/working-group/) in the first instance. The data are currently held in the Outbreak Data Analysis Platform (ODAP, https://odap.ac.uk/). Researchers seeking to access these data are directed to https://www.phosp.org/resource/ for information and forms. Correspondence to be directed to Dr Rachael A Evans, the Co-Principal Investigator of PHOSP-COVID study.

## Data Availability

The PHOSP-COVID study website (https://www.phosp.org) contains an overview of the
study, resources, information about people involved, and publications. Research activity
using the study is organised across a series of Working Groups. These were
established at the outset of the study to coordinate research, minimise duplication of
efforts, and facilitate communication across research and clinical specialties. Researchers
interested in undertaking research using PHOSP-COVID are encouraged to contact the
relevant Working Group leads (https://www.phosp.org/working-group/) in the first
instance. The data are currently held in the Outbreak Data Analysis Platform (ODAP,
https://odap.ac.uk/). Researchers seeking to access these data are directed to
https://www.phosp.org/resource/ for information and forms. Correspondence to be
directed to Dr Rachael A Evans, the Co-Principal Investigator of PHOSP-COVID study.

## Acknowledgements

This study would not be possible without all the participants who have given their time and support. We thank all the participants and their families. We thank the many research administrators, health-care and social-care professionals who contributed to setting up and delivering the study at all of the 65 NHS trusts/Health boards and 25 research institutions across the UK, as well as all the supporting staff at the NIHR Clinical Research Network, Health Research Authority, Research Ethics Committee, Department of Health and Social Care, Public Health Scotland, and Public Health England, and support from the ISARIC Coronavirus Clinical Characterisation Consortium. We thank Kate Holmes at the NIHR Office for Clinical Research Infrastructure (NOCRI) for her support in coordinating the charities group. The PHOSP-COVID industry framework was formed to provide advice and support in commercial discussions, and we thank the Association of the British Pharmaceutical Industry as well NOCRI for coordinating this. We are very grateful to all the charities that have provided insight to the study: Action Pulmonary Fibrosis, Alzheimer’s Research UK, Asthma + Lung UK, British Heart Foundation, Diabetes UK, Cystic Fibrosis Trust, Kidney Research UK, MQ Mental Health, Muscular Dystrophy UK, Stroke Association Blood Cancer UK, McPin Foundations, and Versus Arthritis. We thank the NIHR Leicester Biomedical Research Centre patient and public involvement group and Long Covid Support.

