## Supplementary methods, Table S1 for "One year health outcomes associated with systemic corticosteroids for COVID-19: a longitudinal cohort study"

^1^Department of Population Health Sciences, University of Leicester, Leicester, UK, ^2^The Institute for Lung Health, NIHR Leicester Biomedical Research Centre, University of Leicester, Leicester, UK, ^3^Centre for Medical Informatics, The Usher Institute, University of Edinburgh, Edinburgh, UK, ^4^The Usher Institute, University of Edinburgh, Edinburgh, UK, ^5^Royal Infirmary of Edinburgh, NHS Lothian, Edinburgh, UK, ^6^Roslin Institute, University of Edinburgh, Edinburgh, UK, ^7^University Hospitals of Leicester NHS Trust, Leicester, UK, ^8^Centre for Exercise and Rehabilitation Science, NIHR Leicester Biomedical Research Centre-Respiratory, University of Leicester, Leicester, UK, ^9^Department of Respiratory Sciences, University of Leicester, Leicester, UK, ^10^Therapy Department, University Hospitals of Leicester, NHS Trust, Leicester, UK, ^11^NIHR Leicester Biomedical Research Centre, University of Leicester, Leicester, UK, ^12^St Georges University Hospitals NHS Foundation Trust, London, UK, ^13^The Leeds Teaching Hospitals NHS Trust, Leeds, UK, ^14^University of Nottingham, Nottingham, UK, ^15^Nottingham University Hospitals NHS Trust, Nottingham, UK, ^16^NIHR Nottingham Biomedical Research Centre, Nottingham, UK, ^17^UCL Respiratory, Department of Medicine, University College London, Rayne Institute, London, UK, ^18^University of Edinburgh, Edinburgh, UK, ^19^NHS Lothian, Edinburgh, UK, ^20^Manchester Metropolitan University, Manchester, UK, ^21^Salford Royal NHS Foundation Trust, Manchester, UK, ^22^Infection Research Group, Hull University Teaching Hospitals, Hull, UK, ^23^University of Hull, Hull, UK, ^24^The Newcastle Upon Tyne Hospitals NHS Foundation Trust, Newcastle Upon Tyne, UK, ^25^Translational and Clinical Research Institute, Newcastle University, Newcastle Upon Tyne, UK, ^26^Department of Respiratory Medicine, Cambridge University Hospitals NHS Foundation Trust, Cambridge, UK, ^27^University of Cambridge, Cambridge, UK, ^28^NIHR Cambridge Clinical Research Facility, Cambridge, UK, ^29^Lane Fox Respiratory Service, Guys and St Thomas NHS Foundation Trust, London, UK, ^30^University College London, London, UK, ^31^Royal Free London NHS Foundation Trust, London, UK, ^32^Clinical and Experimental Sciences, Faculty of Medicine, University of Southampton, Southampton, UK, ^33^NIHR Southampton Biomedical Research Centre, University Hospitals Southampton, Southampton, UK, ^34^University of Birmingham, Birmingham, UK, ^35^University Hospital Birmingham NHS Foundation Trust, Birmingham, UK, ^36^Barts Health NHS Trust, London, UK, ^37^Queen Mary University of London, London, UK, ^38^Oxford University Hospitals NHS Foundation Trust, Oxford, UK, ^39^University of Oxford, Oxford, UK, ^40^NIHR Oxford Biomedical Research Centre, Oxford, UK, ^41^CAMS Oxford Institute, Oxford, UK, ^42^University of Sheffield, Sheffield, UK, ^43^Sheffield Teaching NHS Foundation Trust, Sheffield, UK, ^44^Kings College London, London, UK, ^45^Kings College London NHS Foundation Trust, London, UK, ^46^Institute of Infection, Veterinary and Ecological Sciences, University of Liverpool, Liverpool, UK, ^47^Liverpool University Hospitals NHS Foundation Trust, Liverpool, UK, ^48^NIHR Health Protection Research Unit in Emerging and Zoonotic Infections, University of Liverpool, Liverpool, UK, ^49^Department of Psychological Medicine, Institute of Psychiatry, Psychology and Neuroscience, Kings College London, London, UK, ^50^South London and Maudsley NHS Trust, London, UK, ^51^Diabetes Research Centre, University of Leicester, Leicester, UK, ^52^Population Health Sciences Institute, Newcastle University, Newcastle Upon Tyne, UK, ^53^Newcastle upon Tyne Teaching Hospitals Trust, Newcastle upon Tyne, UK, ^54^NIHR Oxford Health Biomedical Research Centre, University of Oxford, Oxford, UK, ^55^Oxford Health NHS Foundation Trust, Oxford, UK, ^56^University of Liverpool, Liverpool, UK, ^57^The CRUK Liverpool Experimental Cancer Medicine Centre, Liverpool, UK, ^58^Department of Oncology and Metabolism, University of Sheffield, Sheffield, UK, ^59^Imperial College Healthcare NHS Trust, London, UK, ^60^Imperial College London, London, UK, ^61^Centre for Medical Image Computing, University College London, London, UK, ^62^Lungs for Living Research Centre, University College London, London, UK, ^63^National Heart and Lung Institute, Imperial College London, London, UK, ^64^MRC-Versus Arthritis Centre for Musculoskeletal Ageing Research, Institute of Inflammation and Ageing, University of Birmingham, Birmingham, UK, ^65^NIHR Birmingham Biomedical Research Centre, University Hospitals Birmingham and the University of Birmingham, Birmingham, UK, ^66^Royal Brompton and Harefield Clinical Group, Guys and St Thomas NHS Foundation Trust, London, UK, ^67^NHLI, Imperial College London, London, UK, ^68^Department of Cardiovascular Sciences, University of Leicester, Leicester, UK, ^69^Division of Cardiovascular Medicine, Radcliffe Department of Medicine, University of Oxford, Oxford, UK, ^70^ILD Service, University College London Hospital, London, UK, ^71^Kadoorie Centre for Critical Care Research, Nuffield Department of Clinical Neurosciences, University of Oxford, Oxford, UK, ^72^MRC-University of Glasgow Centre for Virus Research, Glasgow, UK, ^73^NIHR Health Protection Research Unit in Emerging and Zoonotic Infections, Institute of Infection, Veterinary and Ecological Sciences, University of Liverpool, UK, ^74^Respiratory Medicine, Alder Hey Children’s Hospital, Liverpool, UK, ^75^Cambridge NIHR BRC, Cambridge, UK, ^76^Hywel Dda University Health Board, Wales, UK, ^77^University of Swansea, Wales, UK, ^78^Respiratory Innovation Wales, Wales, UK, ^79^Wellcome-Wolfson Institute for Experimental Medicine, Queens University Belfast, Belfast, UK, ^80^Belfast Health & Social Care Trust, Belfast, UK, ^81^London School of Hygiene & Tropical Medicine, London, UK, ^82^University of Dundee, Ninewells Hospital and Medical School, Dundee, UK, ^83^MRC Human Immunology Unit, University of Oxford, Oxford, UK, ^84^Division of Infection, Immunity & Respiratory Medicine, Faculty of Biology, Medicine and Health, University of Manchester, Manchester, UK, ^85^Manchester University NHS Foundation Trust, Manchester, UK, ^86^Department of Clinical Research, London School of Hygiene & Tropical Medicine, London, UK, ^87^Hospital for Tropical Diseases, University College London Hospital, London, UK, ^88^Division of Infection and Immunity, University College London, London, UK, ^89^Asthma and Lung UK, London, UK, ^90^Radcliffe Department of Medicine, University of Oxford, Oxford, UK

***joint first authors**

**Corresponding author:**

Dr Rachael Evans

NIHR Respiratory BRC, Glenfield Hospital, Groby Rd, Leicester, UK. LE3 9QP

+44 116 258 3663

### PHOSP-COVID Collaborative Group

**Core Management Group**

*Chief Investigator* C E Brightling, *Members* R A Evans (Lead Co-I), L V Wain (Lead Co-I), J D Chalmers, V C Harris, L P Ho, A Horsley, M Marks, K Poinasamy, B Raman, A Shikotra, A Singapuri

**PHOSP-COVID Study Central Coordinating Team**

C E Brightling (Chief Investigator), R A Evans (*Lead Co-I*), L V Wain (*Lead Co-I*), R Dowling, C Edwardson, O Elneima, S Finney, N J Greening, B Hargadon, V C Harris, L Houchen--Wolloff, O C Leavy, H J C McAuley, C Overton, T Plekhanova, R M Saunders, M Sereno, A Singapuri, A Shikotra, C Taylor, S Terry, C Tong, B Zhao

**Steering Committee**

*Co-chairs* D Lomas, E Sapey*, Institution representatives* C Berry, C E Bolton, N Brunskill, E R Chilvers, R Djukanovic, Y Ellis, D Forton, N French, J George, N A Hanley, N Hart, L McGarvey, N Maskell, H McShane, M Parkes, D Peckham, P Pfeffer, A Sayer, A Sheikh, A A R Thompson, N Williams and core management group representation

**Executive Board**

*Chair* C E Brightling, representation from the core management group, each working group and platforms

**Platforms**

**Bioresource**

W Greenhalf (*Co-Lead*), M G Semple (*Co-Lead*), M Ashworth, H E Hardwick, L Lavelle-Langham, W Reynolds, M Sereno, R M Saunders, A Singapuri, V Shaw, A Shikotra, B Venson, L V Wain

**Data Hub**

A B Docherty (*Co-Lead*), E M Harrison (*Co-Lead*), A Sheikh (*Co-Lead*), J K Baillie, C E Brightling, L Daines, R Free, R A Evans, S Kerr, O C Leavy, N I Lone, D Lozano-Rojas, H J C McAuley, K Ntotsis, R Pius, J Quint, M Richardson, , M Sereno, M Thorpe, L V Wain

**Imaging Alliance**

M Halling-Brown (*Co-Lead*), F Gleeson (*Co-Lead*), J Jacob (*Co-Lead*), S Neubauer (*Co-Lead*) B Raman (*Co-Lead*) S Siddiqui (*Co-Lead*) J M Wild (*Co-Lead*), S Aslani, G Baxter, M Beggs, C Bloomfield, M P Cassar, A Chiribiri, E Cox, D J Cuthbertson, M Halling-Brown, V M Ferreira, L Finnigan, S Francis, P Jezzard, G J Kemp, H Lamlum, E Lukaschuk, C Manisty, G P McCann , C McCracken, K McGlynn , R Menke , C A Miller , A J Moss, T E Nichols, C Nikolaidou , C O’Brien , G Ogbole, B Rangelov, D P O’Regan , A Pakzad, S Piechnik , S Plein, I Propescu, A A Samat, L Saunders, Z B Sanders, R Steeds, T Treibel, E M Tunnicliffe, M Webster, J Willoughby, J Weir McCall, C Xie, M Xu

**Omics**

L V Wain (*Co-Lead)*, J K Baillie (*Co-Lead*), H Baxendale, C E Brightling, M Brown, J D Chalmers, R A Evans, B Gooptu, W Greenhalf, H E Hardwick, R G Jenkins, D Jones, I Koychev, C Langenberg, A Lawrie, P L Molyneaux, A Shikotra, J Pearl, M Ralser, N Sattar, R M Saunders, J T Scott, T Shaw, D Thomas, D Wilkinson

**Working Groups**

**Airways**

L G Heaney (*Co-Lead*), A De Soyza (*Co-Lead*), D Adeloye, C E Brightling, J S Brown, J Busby, J D Chalmers, C Echevarria, L Daines, O Elneima, RA Evans, J Hurst, P Novotny, C Nicolaou, P Pfeffer, K Poinasamy, J Quint, I Rudan, E Sapey, M Shankar-Hari, A Sheikh, S Siddiqui, S Walker, B Zheng

**Brain**

J R Geddes (*Lead*), M Hotopf *(Co-Lead),* K Abel, R Ahmed, L Allan, C Armour, D Baguley, D Baldwin, C Ballard, K Bhui, G Breen, K Breeze, M Broome, T Brugha, E Bullmore, D Burn, F Callard, J Cavanagh, T Chalder, D Clark, A David, B Deakin, H Dobson, B Elliott, J Evans, RA Evans, R Francis, E Guthrie, P Harrison, M Henderson,  A Hosseini, N Huneke, M Husain, T Jackson, I Jones, T Kabir, P Kitterick, A Korszun, I Koychev, J Kwan, A Lingford-Hughes, P Mansoori, H McAllister-Williams, K McIvor, B Michael, L Milligan, R Morriss, E Mukaetova-Ladinska, K Munro, A Nevado-Holgado, T Nicholson, C Nicolaou, S Paddick, C Pariante, J Pimm, K Saunders, M Sharpe, G Simons, J P Taylor, R Upthegrove, S Wessely

**Cardiac**

G P McCann (*Lead*), S Amoils, C Antoniades, A Banerjee, A Bularga, C Berry, P Chowienczyk, J P Greenwood, A D Hughes, K Khunti, C Lawson, N L Mills, A J Moss, S Neubauer, B Raman, A N Sattar, C L Sudlow, M Toshner,

**Immunology**

P J M Openshaw (*Lead*), D Altmann, J K Baillie, R Batterham, H Baxendale, N Bishop, C E Brightling, P C Calder, C M Efstathiou, R A Evans, J L Heeney, T Hussell, P Klenerman, F Liew, J M Lord, P Moss, S L Rowland-Jones, W Schwaeble, M G Semple, R S Thwaites, L Turtle, L V Wain, S Walmsley, D Wraith

**Intensive Care**

M J Rowland (*Lead*), A Rostron (*Co-Lead*), J K Baillie, B Connolly, A B Docherty, N I Lone, D F McAuley, D Parekh, A Rostron, J Simpson, C Summers

**Lung Fibrosis**

R G Jenkins (*Co-Lead*), J Porter (*Co-Lead*), R J Allen, R Aul, J K Baillie, S Barratt, P Beirne, J Blaikley, R C Chambers, N Chaudhuri, C Coleman, E Denneny, L Fabbri, P M George, M Gibbons, F Gleeson, B Gooptu, B Guillen Guio, I Hall, N A Hanley, L P Ho, E Hufton, J Jacob, I Jarrold, G Jenkins, S Johnson, M G Jones, S Jones, F Khan, P Mehta, J Mitchell, P L Molyneaux, J E Pearl, K Piper Hanley, K Poinasamy, J Quint, D Parekh, P Rivera-Ortega, L C Saunders, M G Semple, J Simpson, D Smith, M Spears, L G Spencer, S Stanel, I Stewart, A A R Thompson, D Thickett, R Thwaites, L V Wain, S Walker, S Walsh, J M Wild, D G Wootton, L Wright

**Metabolic**

S Heller (*Co-Lead*), M J Davies (*Co-Lead*), H Atkins, S Bain, J Dennis, K Ismail, D Johnston, P Kar, K Khunti, C Langenberg, P McArdle, A McGovern, T Peto, J Petrie, E Robertson, N Sattar, K Shah, J Valabhji, B Young

**Pulmonary and Systematic Vasculature**

L S Howard (*Co-Lead*), Mark Toshner (*Co-Lead*), C Berry, P Chowienczyk, A Lawrie, O C Leavy, J Mitchell, J Newman, L Price, J Quint, A Reddy, J Rossdale, N Sattar, C Sudlow, A A R Thompson, J M Wild, M Wilkins

**Rehabilitation, Sarcopenia and Fatigue**

S J Singh (*Co-Lead*), W D-C Man (*Co-Lead*), J M Lord (*Co-Lead*), N J Greening (*Co-Lead*), T Chalder (*Co-Lead*), J T Scott (*Co-Lead*), N Armstrong, E Baldry, M Baldwin, N Basu, M Beadsworth, L Bishop, C E Bolton, A Briggs, M Buch, G Carson, J Cavanagh, H Chinoy, C Dawson, E Daynes, S Defres, R A Evans, L Gardiner, P Greenhaff, S Greenwood, M Harvie, L HOuchen-Wolloff, M Husain, S MacDonald, A McArdle, H J C McAuley, A McMahon, M McNarry, G Mills, C Nolan, K O’Donnell, D Parekh, Pimm, J Sargent, L Sigfrid, M Steiner, D Stensel, A L Tan, I Vogiatzis, J Whitney, D Wilkinson, D Wilson, M Witham, D G Wootton, T Yates

**Renal**

D Thomas (*Lead*), N Brunskill (*Co-Lead*), S Francis (*Co-Lead*), S Greenwood (*Co-Lead*), C Laing (*Co-Lead*), K Bramham, P Chowdhury, A Frankel, L Lightstone, S McAdoo, K McCafferty, M Ostermann, N Selby, C Sharpe, M Willicombe

**Patient Public Engagement Group**

L Houchen-Wolloff (*Lead*), J Bunker, R Gill, C Hastie, R Nathu, N Rogers, N Smith

**Local Clinical Centre PHOSP-COVID trial staff**

(listed in alphabetical order)

**Airedale NHS Foundation Trust**

A Shaw (PI), L Armstrong, B Hairsine, H Henson, C Kurasz, L Shenton

**Aneurin Bevan University Health Board**

S Fairbairn (PI), A Dell, N Hawkings, J Haworth, M Hoare, A Lucey, V Lewis, G Mallison, H Nassa, C Pennington, A Price, C Price, A Storrie, G Willis, S Young

**Barts Health NHS Trust &** **Queen Mary University of London**

P Pfeffer (PI), K Chong-James, C David, W Y James, C Manisty, A Martineau, O Zongo

**Barnsley Hospital NHS Foundation Trust**

A Sanderson (PI)

**Belfast Health and Social Care Trust & Queen's University Belfast**

L G Heaney (PI), C Armour, V Brown, T Craig, S Drain, B King, N Magee, D McAulay, E Major, L McGarvey, J McGinness, R Stone

**Betsi Cadwaladr University Health Board**

A Haggar (PI), A Bolger, F Davies, J Lewis, A Lloyd, R Manley, E McIvor, D Menzies, K Roberts, W Saxon, D Southern, C Subbe, V Whitehead

**Borders General Hospital, NHS Borders**

H El-Taweel (PI), J Dawson, L Robinson

**Bradford Teaching Hospitals NHS Foundation Trust**

D Saralaya (PI), L Brear, K Regan, K Storton

**Cambridge University Hospitals NHS Foundation Trust, NIHR Cambridge Clinical Research Facility & University of Cambridge**

J Fuld (PI), A Bermperi, I Cruz, K Dempsey, A Elmer, H Jones, S Jose, S Marciniak, M Parkes, C Ribeiro, J Taylor, M Toshner, L Watson, J Weir McCall, J Worsley

**Cardiff and Vale University Health Board**

R Sabit (PI), L Broad, A Buttress, T Evans, M Haynes, L Jones, L Knibbs, A McQueen, C Oliver, K Paradowski, J Williams

**Chesterfield Royal Hospital NHS Trust**

E Harris (PI), C Sampson

**Cwm Taf Morgannwg University Health Board**

C Lynch (PI), E Davies, C Evenden , A Hancock, K Hancock, M Rees , L Roche, N Stroud, T Thomas-Woods

**East Cheshire NHS Trust**

M Babores (PI), J Bradley-Potts, M Holland, N Keenan, S Shashaa, H Wassall

**East Kent Hospitals University NHS Foundation Trust**

E Beranova (PI), H Weston (PI), T Cosier, L Austin, J Deery, T Hazelton, C Price, H Ramos, R Solly, S Turney

**Gateshead NHS Trust**

L Pearce (PI), W McCormick, S Pugmire, W Stoker, A Wilson

**Guy’s and St Thomas’ NHS Foundation Trust**

N Hart (PI), LA Aguilar Jimenez, G Arbane, S Betts, K Bisnauthsing, A Dewar, P Chowdhury, A Chiribiri, A Dewar, G Kaltsakas, H Kerslake, MM Magtoto, P Marino, LM Martinez, C O'Brien, M Ostermann, J Rossdale, TS Solano, E Wynn

**Hampshire Hospitals NHS Foundation Trust**

N Williams (PI), W Storrar (PI), M Alvarez Corral, A Arias, E Bevan, D Griffin, J Martin, J Owen,

S Payne, A Prabhu, A Reed, C Wrey Brown

**Harrogate and District NHD Foundation Trust**

C Lawson (PI), T Burdett, J Featherstone, A Layton, C Mills, L Stephenson,

**Hull University Teaching Hospitals NHS Trust & University of Hull**

N Easom (PI), P Atkin, K Brindle, M G Crooks, K Drury, R Flockton, L Holdsworth, A Richards, D L Sykes, S Thackray-Nocera, C Wright

**Hywel Dda University Health Board**

K E Lewis (PI), A Mohamed (PI), G Ross (PI), S Coetzee, K Davies, R Hughes, R Loosley, L O’Brien, Z Omar, H McGuinness, E Perkins, J Phipps, A Taylor, H Tench, R Wolf-Roberts

**Imperial College Healthcare NHS Trust & Imperial College London**

L S Howard (PI), O Kon (PI), D C Thomas (PI), S Anifowose, L Burden, E Calvelo, B Card, C Carr, E R Chilvers, D Copeland, P Cullinan, P Daly, C M Efstathiou, L Evison, T Fayzan, H Gordon, S Haq, R G Jenkins, C King, F Liew, K March, M Mariveles, L McLeavey, N Mohamed, S Moriera, U Munawar, J Nunag, U Nwanguma, L Orriss- Dib, D P O'Regan, A Ross, M Roy, E Russell, K Samuel, J Schronce, N Simpson, L Tarusan, C Wood, N Yasmin

**Kettering General Hospital NHS Trust**

R Reddy (PI), A-M, Guerdette, M Hewitt, K Warwick, S White

**King’s College Hospital NHS Foundation Trust & Kings College London**

A M Shah (PI), C J Jolley (PI), O Adeyemi, R Adrego, H Assefa-Kebede, J Breeze, M Brown, S Byrne, T Chalder, A Chiribiri, P Dulawan, N Hart, A Hayday, A Hoare, A Knighton, M Malim, C O'Brien, S Patale, I Peralta, N Powell, A Ramos, K Shevket, F Speranza, A Te

**Leeds Teaching Hospitals & University of Leeds**

P Beirne (PI), A Ashworth, J Clarke, C Coupland, M Dalton, E Wade, C Favager, J Greenwood, J Glossop, L Hall, T Hardy, A Humphries, J Murira, D Peckham, S Plein, J Rangeley, G Saalmink, A L Tan, B Whittam, N Window, J Woods,

**Lewisham & Greenwich NHS Trust**

G Coakley (PI)

**Liverpool University Hospitals NHS Foundation Trust & University of Liverpool**

D G Wootton (PI), L Turtle (PI), L Allerton, AM All, M Beadsworth, A Berridge, J Brown, S Cooper, A Cross, D J Cuthbertson, S Defres, S L Dobson, J Earley, N French, W Greenhalf, H E Hardwick, K Hainey, J Hawkes, V Highett, S Kaprowska, G J Kemp, AL Key, S Koprowska, L Lavelle-Langham, N Lewis-Burke, G Madzamba, F Malein, S Marsh, C Mears, L Melling, M J Noonan, L Poll, J Pratt, E Richardson, A Rowe, M G Semple, V Shaw, K A Tripp, B Vinson, L O Wajero, S A Williams-Howard, J Wyles

**London North West University Healthcare NHS Trust**

S N Diwanji (PI), P Papineni (PI), S Gurram, S Quaid, G F Tiongson, E Watson

**Manchester University NHS Foundation Trust & University of Manchester**

B Al-Sheklly (PI), A Horsley (PI), C Avram, P Barran, J Blaikely, M Buch, N Choudhury, D Faluyi, T Felton, T Gorsuch, N A Hanley, T Hussell, Z Kausar, C A Miller, N Odell, R Osbourne, K Piper Hanley, K Radhakrishnan, S Stockdale, D Trivedi

**Newcastle upon Tyne Hospitals NHS Foundation Trust & University of Newcastle**

A De Soyza (PI), C Echevarria (PI), A Ayoub, J Brown, G Burns, G Davies, H Fisher, C Francis, A Greenhalgh, P Hogarth, J Hughes, K Jiwa, G Jones, G MacGowan, D Price, A Sayer, J Simpson, H Tedd, S Thomas, S West, M Witham, S Wright, A Young

**NHS Dumfries and Galloway**

M J McMahon (PI), P Neill

**NHS Greater Glasgow and Clyde Health Board & University of Glasgow**

D Anderson (PI), H Bayes (PI), C Berry (PI), D Grieve (PI), I B McInnes (PI), N Basu, A Brown, A Dougherty, K Fallon, L Gilmour, K Mangion, A Morrow, K Scott, R Sykes, R Touyz

**NHS Highland**

E K Sage (PI), F Barrett, A Donaldson

**NHS Lanarkshire**

M Patel (PI), D Bell, A Brown, M Brown, R Hamil, K Leitch, L Macliver, J Quigley, A Smith, B Welsh

**NHS Lothian & University of Edinburgh**

G Choudhury (PI), J K Baillie, S Clohisey, A Deans, A B Docherty, J Furniss, E M Harrison, S Kelly, N I Lone, D E Newby, A Sheikh

**NHS Tayside & University of Dundee**

J D Chalmers (PI), D Connell, A Elliott, C Deas, J George, S Mohammed, J Rowland, A R Solstice, D Sutherland, C J Tee

**North Bristol NHS Trust & University of Bristol**

N Maskell (PI), D Arnold, S Barrett, H Adamali, A Dipper, S Dunn, A Morley, L Morrison, L Stadon, S Waterson, H Welch

**North Middlesex Hospital NHS Trust**

B Jayaraman (PI), T Light

**Nottingham University Hospitals NHS Trust & University of Nottingham**

C E Bolton (PI), P Almeida, J Bonnington, M Chrystal, E Cox, C Dupont, S Francis, P Greenhaff, A Gupta, L Howard, W Jang, S Linford, L Matthews, R Needham, A Nikolaidis, S Prosper, K Shaw, A K Thomas

**Oxford University Hospitals NHS Foundation Trust & University of Oxford**

L P Ho (PI), N M Rahman (PI), M Ainsworth, A Alamoudi, M Beggs, A Bates, A Bloss, A Burns, P Carter, M Cassar, K M Channon, J Chen, F Conneh, T Dong, R I Evans, E Fraser, X Fu, J R Geddes, F Gleeson, P Harrison, M Havinden-Williams, P Jezzard, N Kanellakis, I Koychev, P Kurupati, X Li, E Lukaschuk, K McGlynn, H McShane, C Megson, K Motohashi, S Neubauer, D Nicoll, G Ogg, E Pacpaco, M Pavlides, Y Peng, N Petousi, J Propescu, N Rahman, B Raman, M J Rowland, K Saunders, M Sharpe, N Talbot, E Tunnicliffe

**Royal Brompton and Harefield Clinical Group, Guy’s and St Thomas’ NHS Foundation Trust.**

W D-C Man (PI), B Patel (PI), R E Barker, D Cristiano, N Dormand, M Gummadi, S Kon, K Liyanage, C M Nolan, S Patel, O Polgar, P Shah, S J Singh, J A Walsh

**Royal Free London NHS Foundation Trust**

J Hurst (PI), H Jarvis (PI), S Mandal (PI), S Ahmad, S Brill, L Lim, D Matila, O Olaosebikan, C Singh

**Royal Papworth Hospital NHS Foundation Trust**

M Toshner (PI), H Baxendale, L Garner, C Johnson, J Mackie, A Michael, J Pack, K Paques, H Parfrey, J Parmar

**Salford Royal NHS Foundation Trust**

N Diar Bakerly (PI), P Dark, D Evans, E Hardy, A Harvey, D Holgate, S Knight, N Mairs, N Majeed, L McMorrow, J Oxton, J Pendlebury, C Summersgill, R Ugwuoke, S Whittaker

**Salisbury NHS Foundation Trust**

W Matimba-Mupaya (PI), S Strong-Sheldrake

**Sheffield Teaching NHS Foundation Trust & University of Sheffield**

S L Rowland-Jones (PI), A A R Thompson (Co PI), J Bagshaw, M Begum, K Birchall, R Butcher, H Carborn, F Chan, K Chapman, Y Cheng, L Chetham, C Clark, Z Coburn, J Cole, M Dixon, A Fairman, J Finnigan, L Finnigan, H Foot, D Foote, A Ford, R Gregory, K Harrington, L Haslam, L Hesselden, J Hockridge, A Holbourn, B Holroyd-Hind, L Holt, A Howell, E Hurditch, F Ilyas, C Jarman, A Lawrie, E Lee, J-H Lee, R Lenagh, A Lye, I Macharia, M Marshall, A Mbuyisa, J McNeill, S Megson, J Meiring, L Milner, S Misra, H Newell, T Newman, C Norman, L Nwafor, D Pattenadk, M Plowright, J Porter, P Ravencroft, C Roddis, J Rodger, P Saunders, J Sidebottom, J Smith, L Smith, N Steele, G Stephens, R Stimpson, B Thamu, N Tinker, K Turner, H Turton, P Wade, S Walker, J Watson, J M Wild, I Wilson, A Zawia

**St George’s University Hospitals NHS Foundation Trust**

R Aul (PI), M Ali, A Dunleavy (PI), D Forton, N Msimanga, M Mencias, T Samakomva, S Siddique, J Teixeira, V Tavoukjian

**Sherwood Forest Hospitals NHS Foundation Trust**

J Hutchinson (PI), L Allsop, K Bennett, P Buckley, M Flynn, M Gill, C Goodwin, M Greatorex, H Gregory, C Heeley, L Holloway, M Holmes, J Kirk, W Lovegrove, TA Sewell, S Shelton, D Sissons, K Slack, S Smith, D Sowter, S Turner, V Whitworth, I Wynter

**Shropshire Community Health NHS Trust**

L Warburton (PI), S Painter, J Tomlinson

**Somerset NHS Foundation Trust**

C Vickers (PI), T Wainwright, D Redwood, J Tilley, S Palmer

**Swansea Bay University Health Board**

G A Davies (PI), L Connor, A Cook, T Rees, F Thaivalappil, C Thomas

**Tameside and Glossop Integrated Care NHS Foundation**

A Butt (PI), M Coulding, H Jones, S Kilroy, J McCormick, J McIntosh, H Savill, V Turner, J Vere

**The Great Western Hospital Foundation Trust**

E Fraile (PI), J Ugoji

**The Hillingdon Hospitals NHS Foundation Trust**

S S Kon (PI), H Lota, G Landers, M Nasseri, S Portukhay

**The Rotherham NHS Foundation Trust**

A Hormis (PI), A Daniels, J Ingham, L Zeidan

**United Lincolnshire Hospitals NHS Trust**

M Chablani (PI), L Osborne

**University College London Hospital & University College London**

M Marks (PI), J S Brown (PI), N Ahwireng, B Bang, D Basire, R C Chambers, A Checkley, R Evans, M Heightman, T Hillman, J Hurst, J Jacob, S Janes, R Jastrub, M Lipman, S Logan, D Lomas, M Merida Morillas, A Pakzad, H Plant, J C Porter, K Roy, E Wall, B Williams, M Xu

**University Hospital Birmingham NHS Foundation Trust & University of Birmingham**

D Parekh (PI), N Ahmad Haider, C Atkin, R Baggott, M Bates, A Botkai, A Casey, B Cooper, J Dasgin, K Draxlbauer, N Gautam, J Hazeldine, T Hiwot, S Holden, K Isaacs, T Jackson, S Johnson, V Kamwa, D Lewis,

J M Lord, S Madathil, C McGhee, K Mcgee, A Neal, A Newton Cox, J Nyaboko, D Parekh, Z Peterkin, H Qureshi, B Rangelov, L Ratcliffe, E Sapey, J Short, T Soulsby, R Steeds, J Stockley, Z Suleiman, T Thompson, M Ventura, S Walder, C Welch, D Wilson, S Yasmin, K P Yip

**University Hospitals of Derby and Burton**

P Beckett (PI) C Dickens, U Nanda

**University Hospitals of Leicester NHS Trust & University of Leicester**

C E Brightling (CI), R A Evans (PI), M Aljaroof, N Armstrong, H Arnold, H Aung, M Bakali, M Bakau, M Baldwin, M Bingham, M Bourne, C Bourne, N Brunskill, P Cairns, L Carr, A Charalambou, C Christie, M J Davies, S Diver, S Edwards, C Edwardson, O Elneima, H Evans, J Finch, S Glover, N Goodman, B Gootpu, N J Greening, K Hadley, P Haldar, B Hargadon, V C Harris, L Houchen-Wolloff, W Ibrahim, L Ingram, K Khunti, A Lea, D Lee, D Lozano-Rojas, G P McCann, H J C McAuley, P McCourt, T Mcnally, G Mills, A Moss, W Monteiro, K Ntotsis, M Pareek, S Parker, A Rowland, A Prickett, I N Qureshi, R Russell, N Samani, M Sereno, M Sharma, A Shikotra, S Siddiqui, A Singapuri, S J Singh, J Skeemer, M Soares, E Stringer, T Thornton, M Tobin, E Turner, L V Wain, T J C Ward, F Woodhead, J Wormleighton, T Yates, A Yousuf,

**University Hospital Southampton NHS Foundation Trust & University of Southampton**

M G Jones (PI), C Childs, R Djukanovic, S Fletcher, M Harvey, E Marouzet, B Marshall, R Samuel, T Sass, T Wallis, H Wheeler

**Whittington Health NHS**

R Dharmagunawardena (PI), E Bright, P Crisp, M Stern

**Wirral University Teaching Hospital**

A Wight (PI), L Bailey, A Reddington

**Wrightington Wigan and Leigh NHS trust**

A Ashish (PI), J Cooper, E Robinson

**Yeovil District Hospital NHS Foundation Trust**

A Broadley (PI)

**York & Scarborough NHS Foundation Trust**

K Howard (PI), L Barman, C Brookes, K Elliott. L Griffiths, Z Guy, D Ionita, H Redfearn, C Sarginson

A Turnbull

**Health and Care Research Wales**

Y Ellis

**London School of Hygiene & Tropical Medicine (LSHTM)**

M Marks, A Briggs

**NIHR Office for Clinical Research Infrastructure**

K Holmes

**Patient Public Involvement Leads**

Asthma UK and British Lung Foundation Partnership - K Poinasamy, S Walker

**Royal Surrey NHS Foundation Trust**

M Halling-Brown

**South London and Maudsley NHS Foundation Trust & Kings College London**

G Breen, M Hotopf

**Swansea University & Swansea Welsh Network**

K Lewis, N Williams

**Supplementary methods – sensitivity analysis**

Sensitivity analyses were performed using the ISARIC (UK) dataset to address selection bias, treatment bias and survivor bias in PHOSP-COVID. First, a propensity score weighting for corticosteroid treatment was developed in the ISARIC (UK) cohort (survivors and non-survivors) using logistic regression with the following covariates: age, sex, obesity status, Index of Multiple Deprivation, WHO Clinical Progression Scale status, and the presence of specific comorbidities (cardiovascular disease, hypertension, chronic pulmonary disease, liver disease, renal disease, chronic neurological disease, malignancy, diabetes and rheumatological diseases).

Second, using the PHOSP-COVID dataset, a prediction model was developed for EQ-5D-5L utility index at one year. Given the distribution of this measure, several modelling approaches were compared: linear regression, random forest, gradient boosted trees, and generalised linear models with gamma distribution (log link), poisson distribution, and zero-inflated poisson distribution. Based on root-mean-square error and r-squared, linear regression was used with the above covariates. A bootstrapping approach was used (1000 replicates) to capture the uncertainty in the prediction model.

Third, we used this model to calculate predicted EQ-5D-5L utility index at one year for those that survived COVID-19 hospitalisation in the ISARIC cohort (1000 estimates per patient). Adults that did not survive hospitalisation were assigned an EQ-5D-5L value of zero. Participants who were in both ISARIC and PHOSP-COVID cohorts were assigned their PHOSP-COVID EQ-5D-5L value.

Fourth, a further sampling step was performed to subsample the 1000 datasets created above down to the sample size of the PHOSP-COVID dataset, to ensure robust standard errors (1000 random samples of each dataset). The resulting 1,000,000 datasets were used to produce an average treatment effect of corticosteroid exposure on EQ-5D-5L utility index weighted by the inverse of propensity for exposure using linear regression and adjusted using the above variable list (double robust). The results were pooled using Rubin’s rules [1].

The sensitivity analysis addressed selection and survivor bias by using the structure of the ISARIC population (assuming the ISARIC population was similar to all hospitalised patients with COVID-19 meeting eligibility criteria to receive corticosteroids). The ISARIC cohort included participants who did not survive hospitalisation with COVID-19. Biased treatment assignment was accounted for by developing a propensity score with corticosteroid as the dependent variable, which was developed in the ISARIC cohort, and was therefore independent of survival status at hospital discharge.

1. Rubin DB. *Multiple Imputation for Nonresponse in Surveys.* New York: John Wiley and Sons; 2004

**Table S1: Baseline characteristics PHOSP cohort (whole cohort, unimputed and unweighted).**
Data are n, n (%), mean (SD) or median (IQR). Percentages are calculated by category after exclusion of missing data for that variable. SD = standard deviation, IQR = interquartile range, SMD = standardised mean difference

| **Characteristic** |  | **Corticosteroids** | **No corticosteroids** | **Total** | **SMD** |
| --- | --- | --- | --- | --- | --- |
| n |  | 1149 | 739 | 1,888 |  |
| Age at admission, years (mean (SD)) |  | 57.95 (12.05) | 59.68 (12.38) | 58.63 (12.21) | 0.141 |
| Sex (%) | Male | 743 (64.7) | 472 (63.9) | 1215 (64.4) | 0.017 |
|  | Female | 406 (35.3) | 267 (36.1) | 673 (35.6) |  |
| Ethnicity (%) | White | 873 (76.8) | 534 (72.5) | 1407 (75.1) | 0.108 |
|  | South Asian | 108 (9.5) | 82 (11.1) | 190 (10.1) |  |
|  | Black | 73 (6.4) | 63 (8.5) | 136 (7.3) |  |
|  | Other | 83 (7.3) | 58 (7.9) | 141 (7.5) |  |
|  | Missing | 12 | < 5 |  |  |
| Index of Multiple Deprivation (%) | 1 - most deprived | 269 (23.6) | 158 (21.6) | 427 (22.8) | 0.193 |
|  | 2 | 296 (25.9) | 142 (19.4) | 438 (23.4) |  |
|  | 3 | 190 (16.7) | 141 (19.2) | 331 (17.7) |  |
|  | 4 | 190 (16.7) | 132 (18.0) | 322 (17.2) |  |
|  | 5 - least deprived | 196 (17.2) | 160 (21.8) | 356 (19.0) |  |
|  | Missing | 8 | 6 | 14 |  |
| Obesity (%) | Yes | 423 (61.0) | 335 (55.8) | 758 (58.6) | 0.106 |
|  | No | 270 (39.0) | 265 (44.2) | 535 (41.4) |  |
|  | Missing | 456 | 139 | 595 |  |
| Smoking history (%) | Never | 553 (54.9) | 334 (56.4) | 887 (55.4) | 0.134 |
|  | Ex-smoker | 432 (42.9) | 254 (42.9) | 686 (42.9) |  |
|  | Current smoker | 23 (2.3) | < 5 | 27 (1.7) |  |
|  | Missing | 141 | 147 | 288 |  |
| Number of comorbidities (median [IQR]) |  | 1.00 [0.00, 2.00] | 1.00 [0.00, 2.00] | 1.00 [0.00, 2.00] | 0.056 |
| Number of comorbidities (%) | No comorbidity | 341 (29.7) | 219 (29.6) | 560 (29.7) | 0.069 |
|  | 1 comorbidity | 318 (27.7) | 184 (24.9) | 502 (26.6) |  |
|  | 2+ comorbidities | 490 (42.6) | 336 (45.5) | 826 (43.8) |  |
| Cardiac comorbidity (%) | Yes | 557 (48.6) | 370 (50.3) | 927 (49.3) | 0.034 |
|  | No | 588 (51.4) | 365 (49.7) | 953 (50.7) |  |
|  | Missing | <5 | < 5 |  |  |
| Metabolic/endocrine/renal comorbidity (%) | Yes | 314 (27.4) | 209 (28.3) | 523 (27.7) | 0.022 |
|  | No | 834 (72.6) | 529 (71.7) | 1363 (72.3) |  |
|  | Missing | < 5 | < 5 |  |  |
| Respiratory comorbidity (%) | Yes | 296 (25.8) | 182 (24.6) | 478 (25.3) | 0.026 |
|  | No | 853 (74.2) | 557 (75.4) | 1410 (74.7) |  |
| Type 2 diabetes (%) | Yes | 229 (19.9) | 167 (22.6) | 396 (21.0) | 0.066 |
|  | No | 919 (80.1) | 571 (77.4) | 1490 (79.0) |  |
|  | Missing | < 5 | < 5 |  |  |
| Neurology/psychiatry comorbidity (%) | Yes | 57 (5.0) | 30 (4.1) | 87 (4.6) | 0.044 |
|  | No | 1090 (95.0) | 708 (95.9) | 1798 (95.4) |  |
|  | Missing | < 5 | < 5 |  |  |
| Respiratory support (%) | WHO scale 5 | 592 (51.5) | 404 (54.7) | 996 (52.8) | 0.274 |
|  | WHO scale 6 | 383 (33.3) | 167 (22.6) | 550 (29.1) |  |
|  | WHO scale 7-9 | 174 (15.1) | 168 (22.7) | 342 (18.1) |  |
